# What do men who have sex with men think of the use of antibiotics as pre- and post-exposure prophylaxis to prevent sexually transmitted infections?

**DOI:** 10.1101/2023.09.06.23295017

**Authors:** Amy Matser, Bas Hulstein, Henry J.C. de Vries, Elske Hoornenborg, Maria Prins, Udi Davidovich, Maarten Schim van der Loeff

**Affiliations:** Department of infectious diseases, Public Health Service of Amsterdam, Amsterdam, the Netherlands; Amsterdam UMC location University of Amsterdam, Department of Internal Medicine, Meibergdreef 9, Amsterdam, The Netherlands; Amsterdam Institute for Infection and Immunity (AII), Infectious Diseases, Amsterdam, the Netherlands; Amsterdam Public Health Research Institute (APH), Amsterdam, the Netherlands; Department of Dermatology, Amsterdam UMC, Amsterdam, the Netherlands; Amsterdam Institute for Global Health and Development, Amsterdam, the Netherlands

## Abstract

**OBJECTIVE:** Clinical trials have shown that doxycycline as post-exposure prophylaxis after sexual contact (doxy-PEP) prevents sexually transmitted bacterial infections (STI). We investigated current awareness about informal use of antibiotics as pre- and post-exposure prophylaxis to prevent STI (STI-PrEP/PEP) among men who have sex with men (MSM). In addition, we investigated psychosocial determinants of its use.

**METHODS:** Data were collected in the Amsterdam Cohort Study among MSM, the Netherlands, between October 2021 and October 2022. In an online questionnaire, we assessed socio-demographics, sexual behavior, bacterial STI diagnoses, STI-PrEP/PEP awareness, perceived effectiveness of, and beliefs and attitudes towards STI-PrEP/PEP, and intention to use it. STI-PrEP/PEP users were described and (ordinal) logistic regression analysis was conducted to identify factors associated with STI-PrEP/PEP awareness (yes/no) and intention to use STI-PrEP/PEP (7-point Likert scale).

**RESULTS:** Among 593 MSM with median age 46 years (IQR 36-53), 102 (17.2%) were aware of STI-PrEP/PEP and 15 (2.5%) had ever used it. STI-PrEP/PEP awareness was associated with living with HIV, HIV-PrEP use in the preceding 6 months, and sexualized drug use with casual partner(s). Median intention to use STI-PrEP-PEP was 3 (IQR 2-4). Higher intention to use STI-PrEP/PEP was associated with HIV-PrEP use, sexual contact with casual partners, being worried to get an STI, self-protection as reason to use it, the intention to reduce STI testing and sexual experimenting. Stigmatizing beliefs regarding STI-PrEP/PEP users were associated with lower use intentions.

**CONCLUSION:** Preventive use of antibiotics for STI prevention is limited among MSM in the Netherlands in 2021/2022. Some men have a high intention for future use. Self-protection and a wish for sexual experimenting are amongst the intrinsic motivators for higher intention to use STI-PrEP/PEP. More studies on the safe use of STI-PrEP/PEP are required as well as a strategy to educate those who have already adopted STI-PrEP/PEP or have a high intention to do so, and their healthcare providers.

## INTRODUCTION

In the last two decades, highly effective biomedical prevention strategies for HIV have been developed, such as Treatment as Prevention (TasP) of persons living with HIV, and the use of pre-exposure prophylaxis (HIV-PrEP) for persons who have a higher probability of acquiring HIV(1,2). The implementation of TasP and HIV-PrEP among men who have sex with men (MSM) in high-income countries is associated with a decreasing number of HIV infections. At the same time we have seen a decoupling of declining HIV numbers on the one hand, and rising number of other STI on the other hand (3–5). The rise of other STI predates the widespread implementation of biomedical HIV preventive measures such as TasP and PrEP, and is not fully explained yet, although PrEP use has been associated with decreasing condom use and increasing STI(6,7).

Recently, biomedical prevention of bacterial STI has been introduced as a potential strategy to curb STI spread. The scientific debate on the use of doxycycline as post-exposure prophylaxis (doxy-PEP) gained momentum in 2018 when Molina et al. published the results on the efficacy of doxy-PEP to prevent bacterial STI among MSM participating in the ANRS IPERGAY trial, a French double-blind study on the efficacy of on-demand PrEP for HIV (8). Results from this and two other clinical studies done in the United States showed that the use of doxy-PEP within 72 hours after sexual contact effectively reduces the probability of developing chlamydiasis and syphilis (8–10). In the French IPERGAY trial, doxy-PEP was not effective against gonorrhea, probably due to the high prevalence of tetracycline antimicrobial resistant (AMR) *N. gonorrhoeae* strains (8). In contrast, a doxy-PEP study in Seattle and San Francisco did prove effectivity against gonorrhea, probably due to different antimicrobial resistance (AMR) patterns between sites. Based on these trial results, in October 2022, the San Francisco Department of Public Health published an online health update that recommends the use of doxy-PEP for cis men and trans women who have had a syphilis diagnosis or a bacterial STI and who also report condomless anal or oral sex with a cis male or trans woman in the past year (11). However, no other countries or professional bodies recommend use of doxy-PEP to prevent STI.

Use of doxy-PEP seems paradoxical, because it contravenes the idea of antimicrobial stewardship and may increase AMR (12), particularly in the Netherlands where antibiotic use is relatively low (13,14). Yet, if doxy-PEP could prevent infections that otherwise require antibiotic treatment, it may lead to a reduction of the total amount of antibiotics used and may limit use of certain classes of antibiotics deemed critical for indications hampered by emerging AMR. Doxycycline is already widely used chronically for its anti-inflammatory action in the treatment of inflammatory skin disease such as rosacea and acne, and to prevent malaria (15,16). Moreover, doxycycline and other tetracyclines are not considered “drugs of last resort” for the treatment of infections caused by multi-resistant bacteria.

Prevention strategies are only effective when the key population is willing to adopt the strategy. According to social science theories, such as the Theory of Planned Behavior, having more positive beliefs and attitudes increase intention an actual behavior (17). A mixed method study in 2011 found that chemoprophylaxis to prevent syphilis was likely to be an acceptable intervention for syphilis prevention among men who have sex with men (MSM)(5). In addition, anecdotal evidence from our Center for Sexual Health (CSH) in Amsterdam, the Netherlands, shows that some visitors (mainly MSM), already informally use antibiotics as pre- or post-exposure prophylaxis to prevent STI (STI-PrEP/PEP). This study was performed to gain insight into current informal use, awareness of STI-PrEP/PEP as an STI prevention strategy, and beliefs, attitudes and intention towards STI-PrEP/PEP use among MSM in Amsterdam, the Netherlands.

## METHODS

The study was performed in the Amsterdam Cohort Study among MSM (ACS), which is an open, prospective cohort study initiated in 1984. The study includes HIV-negative MSM and MSM living with HIV. The ACS aims to investigate the epidemiology, natural history, and pathogenesis of HIV and other STI, and to evaluate the effect of interventions (3,18). Inclusion criteria are: being male, being at least 16 years of age, living in Amsterdam or having a strong connection to the city, and having had sexual contact with a man in the preceding 6 months. Biannually, participants complete an online questionnaire about health and sexual behavior and they visit the Public Health Service of Amsterdam for HIV and other STI testing. Participants who are using HIV-PrEP have two additional visits per year, at which HIV/STI tests are performed, in agreement with the Dutch PrEP guidelines (19). In 2022, the ACS was re-approved by the Medical Ethics Committee of the Amsterdam UMC, the Netherlands (MEC 2007-182, NL18679.018.07).

### Data collection

For this study, we used data collected between October 2021 and October 2022, on the informal use of, and awareness, beliefs, attitudes and intention towards the use of STI-PrEP/PEP. We selected the first study visit with complete questionnaire data. Participants, irrespective of HIV status or HIV-PrEP use, were included in this analysis.

### Laboratory methods

HIV testing was done for HIV-1 using HIVAg/Ab combo tests (LIAISON XL Murex; Diasorin, Saluggia, Italy). A western blot (HIV Blot 2.2; Diasorin) and an HIV-1 RNA load test (Abbott RealTime HIV-1; Abbott Molecular Diagnostics B.V., Hoofddorp, the Netherlands) were performed if an HIV Ag/Ab combo test was positive or indeterminate. STI tests included a *Treponema pallidum* particle agglutination assay for detection of *T. pallidum* (syphilis) (LIAISONXL; Diasorin). To diagnose and confirm syphilis infection, the rapid plasma reagin card test and the fluorescent treponemal antibody (FTA)-absorption test (Nosticon and Trepo-spot IF; Biom_erieux, Marcy l’Etoile, France) were performed. Nucleic acid amplification tests were used to detect infection with *Neisseria gonorrhoeae* (gonorrhea) or *Chlamydia trachomatis* (chlamydia) (Aptima Combo 2; Hologic, Marlborough, Massachusetts, USA).

### Variables

Sociodemographic characteristics included in the analysis were age in years, country of birth categorized as the Netherlands or other country, and education categorized as high vocational training/university or other education. Also included were HIV-status and HIV-PrEP use in the preceding six months. Sexual behavior variables were having a sexual relationship with a steady partner, having casual known and anonymous partner(s), anal sex without condoms and oral sex with these casual partners, and group sex, all in the preceding 6 months. Sexualized drug use was defined as the use of mephedrone, methamphetamine, gamma-hydroxybutyric acid/gamma-butyrolactone, ketamine, amphetamines, cocaine, XTC/MDMA, psychedelics, or benzodiazepines during sex with casual partners in the preceding 6 months (7). Travelling to a foreign country in the preceding 6 months was also considered, as was the number of visits to the CSH in the preceding 6 months at which syphilis, chlamydia and/or gonorrhea was diagnosed. We collected data on perceptions of past risk, severity and attitudes towards STI’s by asking how likely the participants estimated that they had contracted these STI in the past 6 months, how scared they had been to contract an STI and how important it had been for them to prevent these infections, all measured on a 7-point Likert scale.

Participants were asked whether they were aware or not of antibiotics use as pre-exposure and post-exposure prophylaxis to prevent STI. Participants were not given any additional information on effectiveness, side-effects, costs, or other aspects of STI-PrEP/PEP before answering questions about beliefs and attitudes towards STI-PrEP/PEP use. The intention to use STI-PrEP/PEP was assessed using the following two questions: “How likely are you to use antibiotics as PrEP or PEP for STI if it becomes available in the Netherlands” and “Are you planning on using antibiotics as PrEP or PEP for STI if it becomes available in the Netherlands”, both measured on a 7-point Likert scale ranging from ‘very unlikely’ (1) to ‘very likely’ (7). Given the high correlation (Cronbach’s α = 0.95) the mean score of these items was used to represent ‘intention to use antibiotics as PrEP or PEP for STI’. High intent was defined as score 6-7. Moreover, participants were asked about the use of STI PrEP/PEP in the past six months: “Have you used antibiotics as PrEP or PEP for STI in the past six months?”

### Statistical analysis

First, characteristics of individuals who were aware of STI-PrEP/PEP versus those who were not were described. Univariable and multivariable logistic regression analyses were done to determine factors associated with STI-PrEP/PEP awareness. Second, characteristics of STI-PrEP/PEP users were compared with those of non-users using Pearson’s chi-squared tests for categorical variables and Kruskal-Wallis tests for continuous variables. Third, factors associated with intention to use STI-PrEP/PEP were determined using univariable and multivariable ordinal logistic regression analysis. Factors for multivariable analysis were selected based on p-value cut-off of p<0.200. A stepwise backward selection method was used to determine the final multivariable model. P<0.050 was considered statistically significant. Analyses were done in STATA 15.

## RESULTS

We included 593 MSM who completed at least one questionnaire in the study period. The median age of the participants was 46 years (IQR 36-53), 484 (86.3%) MSM were born in the Netherlands, 466 (80.2%) had a high vocational or university degree, 36 (6.1%) were living with HIV, and 234 (42.1%) MSM reported HIV-PrEP use in the preceding 6 months.

Sexual contact with a steady partner in the preceding 6 months was reported by 306 (51.6%) MSM. Casual partners were reported by 414 (69.9%) MSM. Half of the participants reported condomless anal sex (292; 49.3%), oral sex (412; 69.6%) or anilingus (313; 52.9%) in the preceding 6 months. Group sex was reported by 414 (69.9%) individuals and sexualized drug use with a casual partner by 156 (26.4%) individuals. In total, 25 (4.3%) individuals were diagnosed with chlamydia, 31 (5.3%) with gonorrhea, and 6 (1.0%) with syphilis.

### STI-PrEP/PEP awareness

In total, 102 (17.2%) participants were aware of STI-PrEP/PEP as prevention strategy, 42 (7.4%) reported at least one casual sex partner who had used STI-PrEP/PEP and 6 (1.1%) had a steady partner who had used it. In univariable analysis, individuals who were aware of STI-PrEP/PEP were more often living with HIV or more likely to have used HIV-PrEP in the preceding 6 months. They more often reported sex with casual known partners, sexualized drug use, group sex, condomless anal and oral sex, anilingus, and they were more likely to have been diagnosed with an STI in the preceding 6 months (Table 1). In multivariable analysis, being aware of STI-PrEP/PEP remained significantly associated with living with HIV (adjusted odds ratio [aOR] 2.82; 95% confidence interval [CI] 1.28-6.23), PrEP use in the preceding 6 months (aOR 1.67; 95% CI 1.03-2.72), and sexualized drug use (aOR 1.89; 95% CI 1.17-3.05) (Table 1).

**Table 1.**
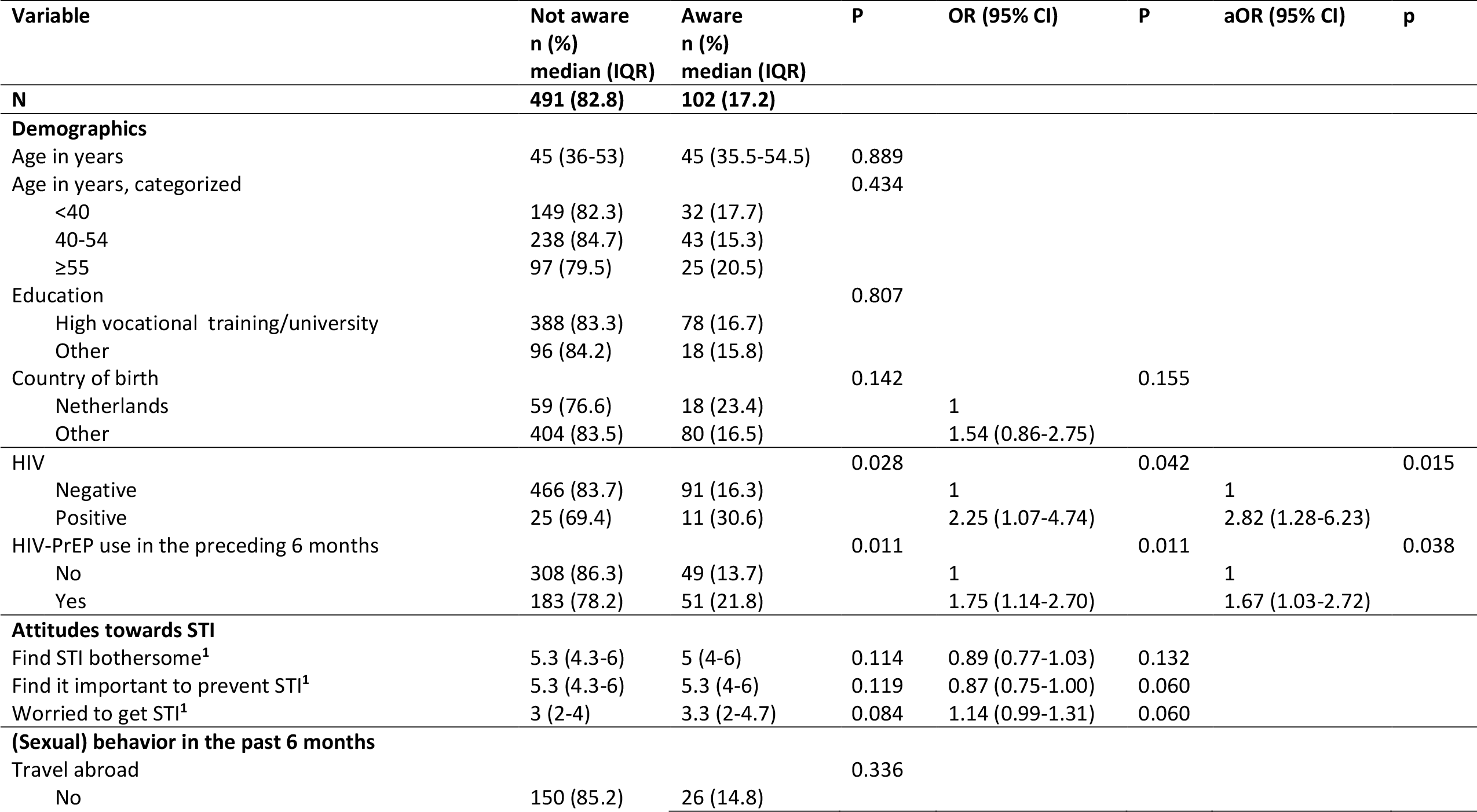
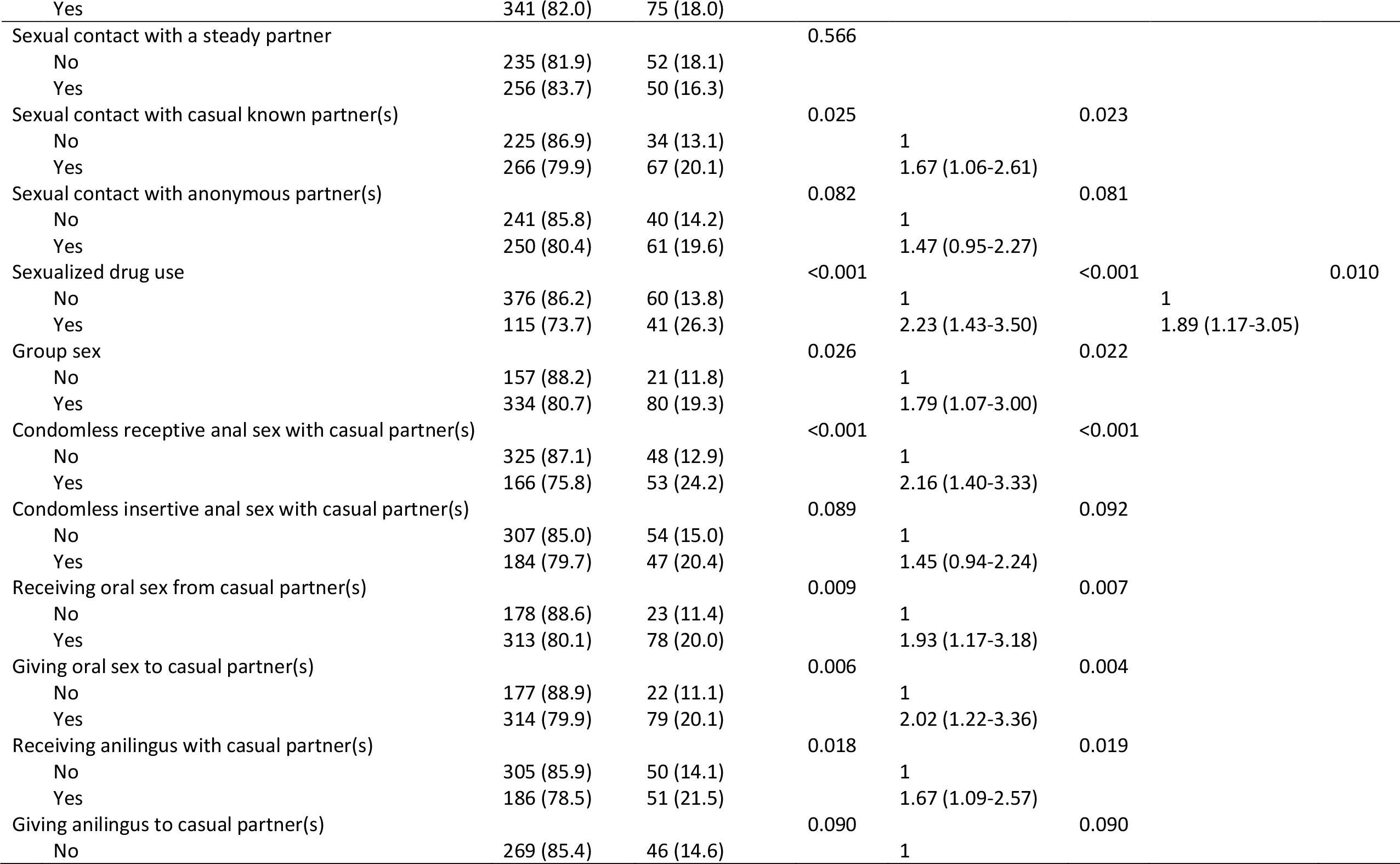
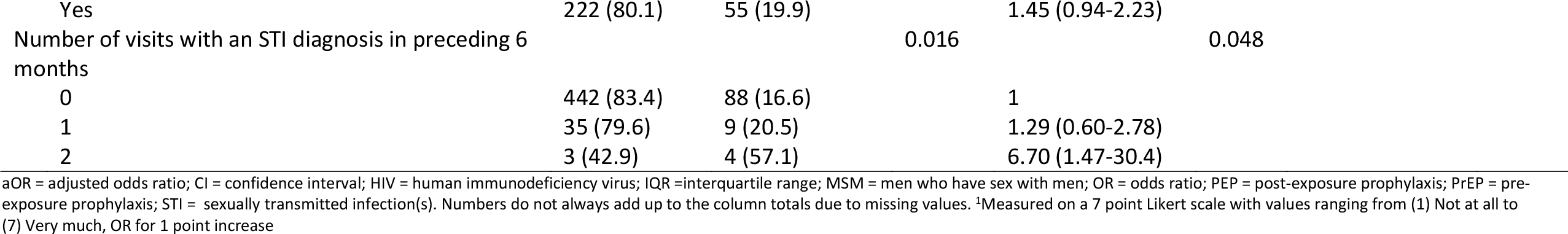
Population characteristics and factors associated with awareness of STI-PrEP/PEP resulting from logistic regression analysis among 593 MSM participating in the Amsterdam Cohort Study on HIV, the Netherlands, October 2021-October 2022.

### STI-PrEP/PEP use

Only 15 (2.5%) individuals had ever used STI-PrEP/PEP. Individuals reported that they had obtained STI-PrEP/PEP via a steady partner (n=2), friends (n=2), online (n=4), abroad (n=3), a scientific study (n=2), or a physician (n=2). Six MSM reported the use of doxycycline, 2 of azithromycin, 1 of erythromycin, 4 reported unknown/other antibiotics, and data was missing for 2 MSM. Ten MSM had used STI-PrEP/PEP in the preceding 6 months of whom 6 had used in once per month or less, 3 had used it 2 to 4 times a month, and 1 had used it 4 times a week or more. At the last episode, 5 had used in for just 1 day, 1 had used it for the 3 consecutive days, 1 for 5 days, and 1 for 7 days. STI-PrEP/PEP use was more common among MSM who had used HIV-PrEP in the preceding 6 months, compared to those not on HIV-PrEP (i.e. 4.7% vs. 1.1%; p=0.007) and among MSM who reported sexualized drug use (i.e. 6.4% vs. 1.2%; p<0.001) (Table 2). Among those who reported receptive condomless anal sex (rCAS) with casual partners, 4.6% had used STI-PrEP/PEP, while this percentage was 1.3% among those who did not report rCAS (p=0.016). The number of MSM with partners who had used STI-PrEP/PEP was low, but there was an association between own use and use by the partner(s): 66.7% of STI-PrEP/PEP users and 5.5% of non-users reported a partner who had used STI-PrEP/PEP (p<0.001).

**Table 2.** The characteristics of individuals who never and those who had ever used STI-PEP/PrEP among 593 MSM participating in the Amsterdam Cohort Study on HIV, the Netherlands, October 2021-October 2022.

| Variable | Never used<br>n (%)<br>median<br>(IQR) | Ever used<br>n (%)<br>median<br>(IQR) | P |
| --- | --- | --- | --- |
| **N** | **578 (97.5)** | **15 (2.5)** |  |
| **Demographics** |  |  |  |
| Age in years | 46 (36-53) | 42 (34-57) | 0.865 |
| Age in years, categorized |  |  | 0.037 |
| <40 | 174 (96.1) | 7 (3.9) |  |
| 40-54 | 279 (99.3) | 2 (0.7) |  |
| ≥55 | 117 (95.9) | 5 (4.1) |  |
| Education |  |  | 0.973 |
| High vocational training/university | 454 (97.4) | 12 (2.6) |  |
| Other | 111 (97.4) | 3 (2.6) |  |
| Country of birth |  |  | 0.474 |
| Netherlands | 74 (96.1) | 3 (3.9) |  |
| Other | 472 (97.5) | 12 (2.5) |  |
| HIV |  |  | 0.922 |
| Negative | 543 (97.5) | 14 (2.5) |  |
| Positive | 35 (97.2) | 1 (2.8) |  |
| HIV-PrEP use in the preceding 6 months |  |  | 0.007 |
| No | 353 (98.9) | 4 (1.1) |  |
| Yes | 223 (95.3) | 11 (4.7) |  |
| **Attitudes towards STI** |  |  |  |
| Find STI bothersome<sup>1</sup> | 5.3 (4-6) | 5.3 (5-6.3) | 0.497 |
| Find it important to prevent STI<sup>1</sup> | 5.3 (4.3-6.3) | 5.3 (4.3-6.3) | 0.687 |
| Worried to get STI<sup>1</sup> | 3 (2-4.3) | 4 (2-4.7) | 0.314 |
| **(Sexual) behavior in the past 6 months** |  |  |  |
| Travel abroad |  |  | 0.793 |
| No | 172 (97.7) | 4 (2.3) |  |
| Yes | 405 (97.4) | 11 (2.6) |  |
| Sexual contact with a steady partner |  |  | 0.237 |
| No | 282 (98.3) | 5 (1.7) |  |
| Yes | 296 (96.7) | 10 (3.3) |  |
| Sexual contact with casual known partner(s) |  |  | 0.410 |
| No | 254 (98.1) | 5 (1.9) |  |
| Yes | 323 (97.0) | 10 (3.0) |  |
| Sexual contact with anonymous partner(s) |  |  | 0.267 |
| No | 276 (98.2) | 5 (1.8) |  |
| Yes | 301 (96.8) | 10 (3.2) |  |
| Sexualized drug use |  |  | <0.001 |
| No | 431 (98.9) | 5 (1.2) |  |
| Yes | 146 (93.6) | 10 (6.4) |  |
| Group sex |  |  | 0.152 |
| No | 176 (98.9) | 2 (1.1) |  |
| Yes | 401 (96.9) | 13 (3.1) |  |
| Condomless receptive anal sex with casual partner(s) |  |  | 0.016 |
| No | 368 (98.7) | 5 (1.3) |  |
| Yes | 209 (95.4) | 10 (4.6) |  |
| Condomless insertive anal sex with casual partner(s) |  |  | 0.250 |
| No | 354 (98.1) | 7 (1.9) |  |
| Yes | 223 (96.5) | 8 (3.5) |  |
| Receiving oral sex from casual partner(s) |  |  | 0.088 |
| No | 199 (99.0) | 2 (1.0) |  |
| Yes | 378 (96.7) | 13 (3.3) |  |
| Providing oral sex to casual partner(s) |  |  | 0.092 |
| No | 197 (99.0) | 2 (1.0) |  |
| Yes | 380 (96.7) | 13 (3.3) |  |
| Receiving anilingus with casual partner(s) |  |  | 0.037 |
| No | 311 (98.7) | 4 (1.3) |  |
| Yes | 266 (96.0) | 11 (4.0) |  |
| Providing anilingus with casual partner(s) |  |  | 0.001 |
| No | 352 (99.2) | 3 (0.9) |  |
| Yes | 225 (94.9) | 12 (5.1) |  |
| Number of visits with an STI diagnosis in preceding 6 months |  |  | 0.024 |
| 0 | 519 (97.9) | 11 (2.1) |  |
| 1 | 41 (93.2) | 3 (6.8) |  |
| 2 | 6 (85.7) | 1 (14.3) |  |
| Steady partner uses STI-PrEP/PEP |  |  | <0.001 |
| No/don't know | 576 (98.1) | 11 (1.9) |  |
| Yes | 2 (33.3) | 4 (66.7) |  |
| Casual partner(s) use(s) STI-PrEP/PEP |  |  | <0.001 |
| No/don't know | 545 (99.1) | 5 (0.9) |  |
| Yes | 32 (76.2) | 10 (23.8) |  |

HIV = human immunodeficiency virus; IQR =interquartile range; MSM = men who have sex with men; PEP = post-exposure prophylaxis; PrEP = pre-exposure prophylaxis; STI = sexually transmitted infection(s). Numbers do not always add up to the column totals due to missing values. <sup>1</sup>Measured on a 7 point Likert scale with values ranging from (1) Not at all to (7) Very much

### Intention to use STI-PrEP/PEP

In this analysis, 524 MSM were included with detailed data on beliefs and attitudes towards STI-PrEP-PEP use. The median intention to use STI-PrEP/PEP was 3 (IQR 2-4) on a 1-7 Likert scale; 62 (11.8%) reported high intent to use STI/PrEP-PEP. In univariable analysis, HIV-PrEP use, sexual behaviors known to be associated with STI infection and sexualized drug use were significantly associated with a higher intention to use STI-PrEP/PEP (Table 3). Being worried to get an STI, importance of STI (self)-protection as reason to use STI-PrEP-PEP, and the desire for higher sexual pleasure were associated as well. In multivariable analysis, individuals who had used HIV-PrEP in the preceding 6 months and who had had sexual contact with casual partners were more likely to have a higher intention to use STI-PrEP/PEP (aOR 1.58; 95% CI 1.12-2.25 and aOR 1.65; 95% CI 1.16-2.34, respectively). A higher intention to use STI-PrEP/PEP was also significantly associated with being more worried about acquiring an STI (aOR 1.35; 95% CI 1.20-1.52). Reasons to use STI-PrEP/PEP associated with the intention to use STI-PrEP/PEP were the desire of self-protection against STI (aOR 1.55; 95% CI 1.38-1.73), being able to reduce STI testing (aOR 1.28; 95% CI 1.14-1.42), and the desire of sexual experimenting (aOR 1.31; 95% CI 1.18-1.46). MSM who thought that STI-PrEP/PEP users were promiscuous were more likely to have a lower intention to use it (aOR 0.82; 95% CI 0.72-0.92).

**Table 3.**
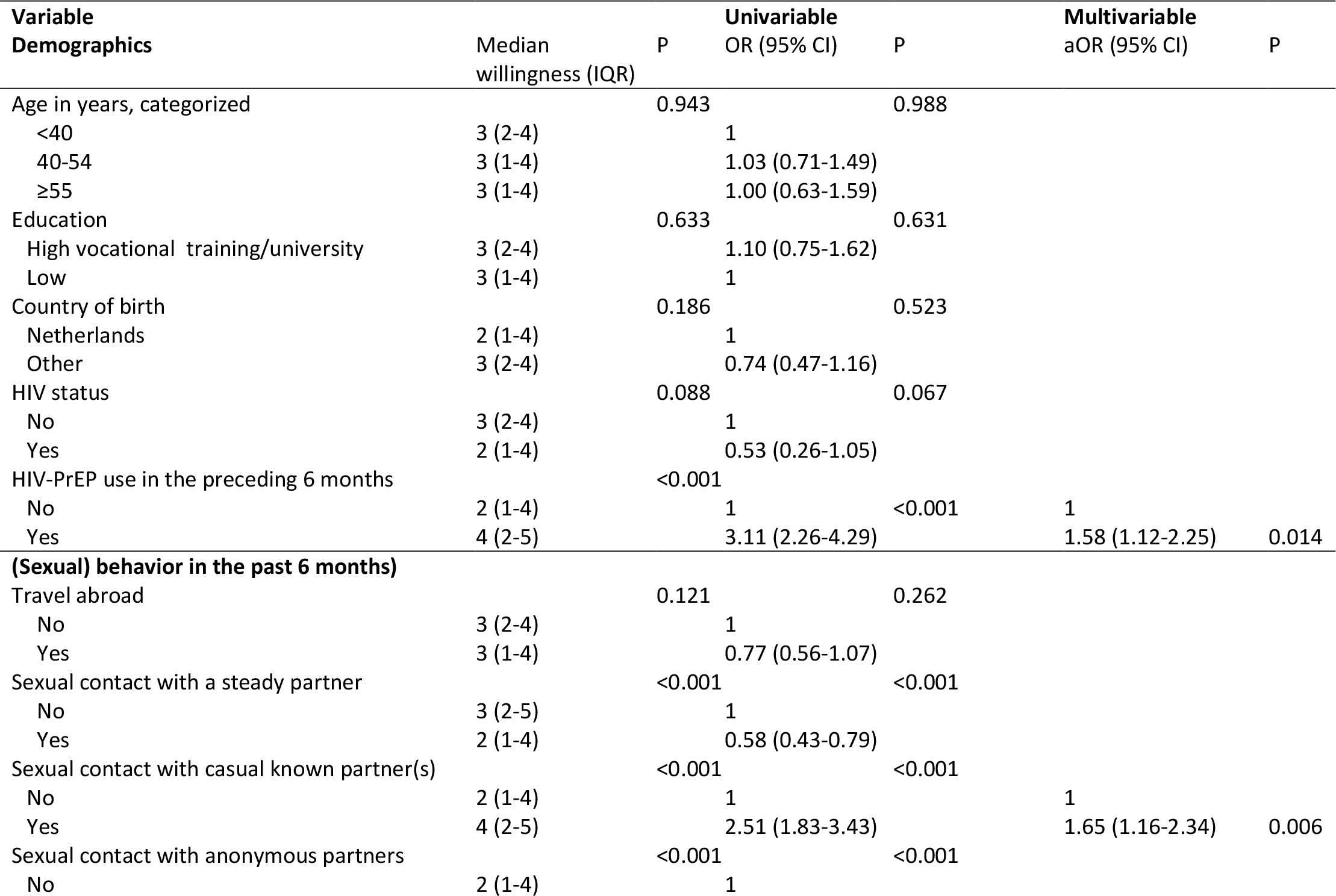
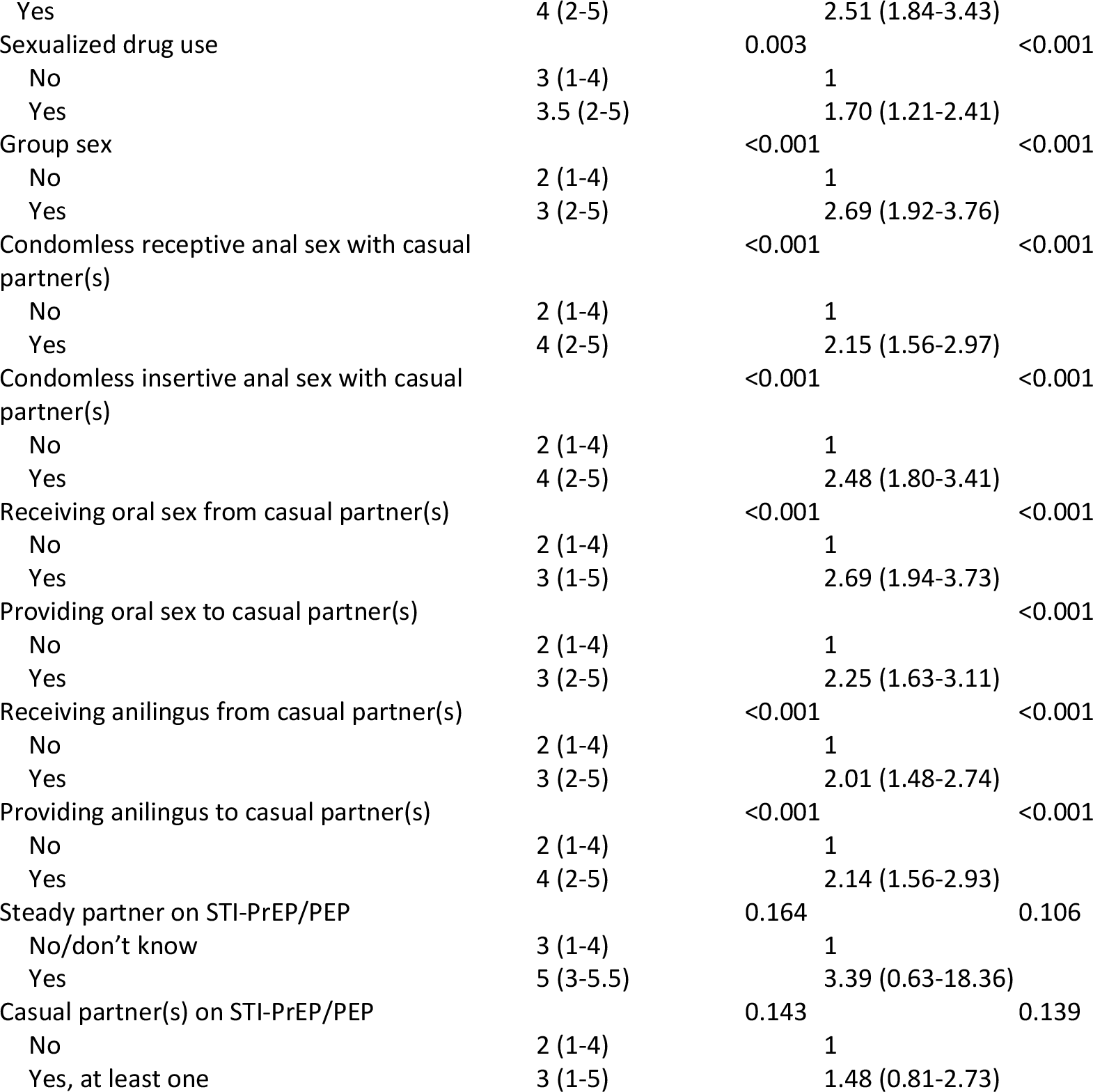
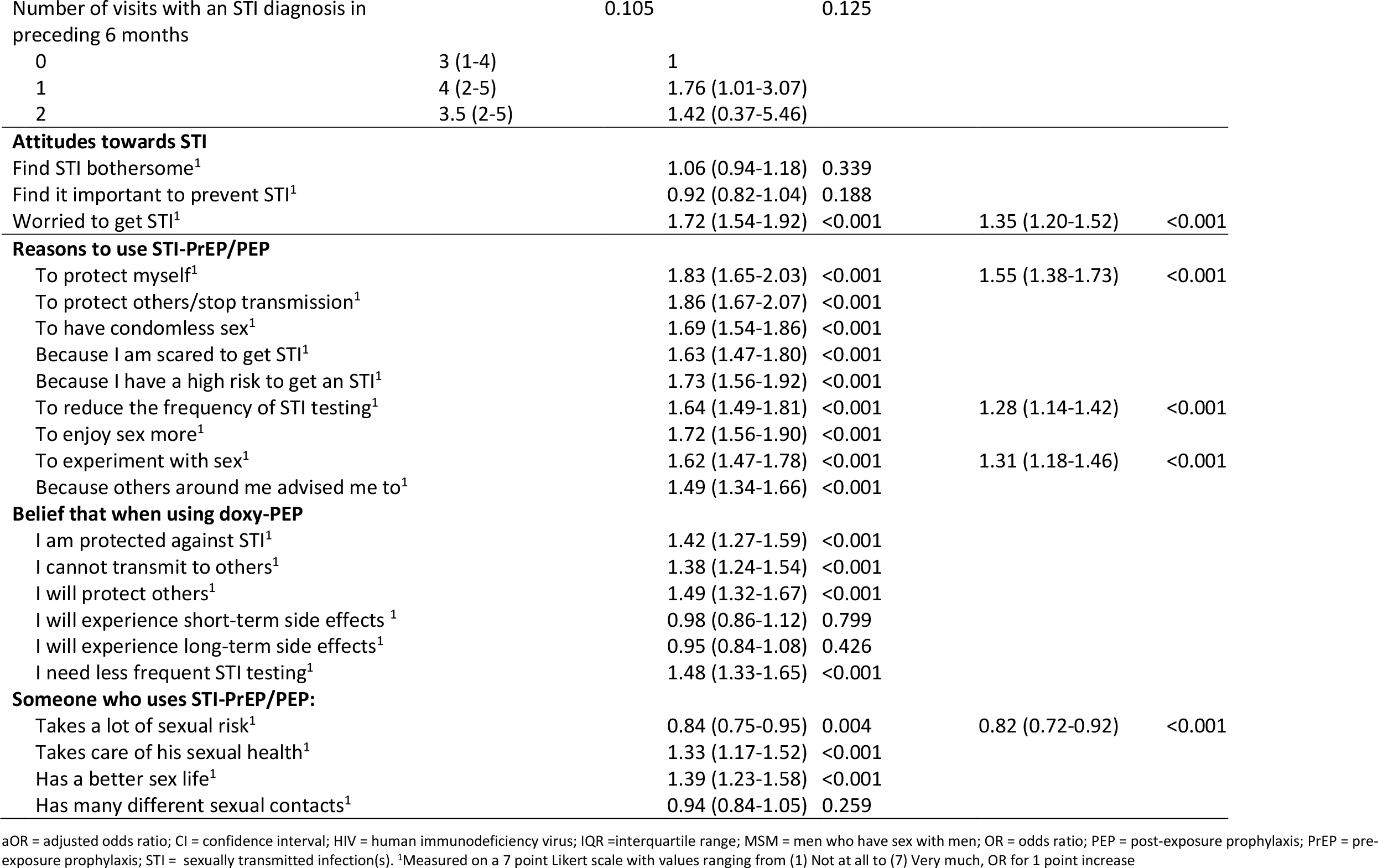
Factors associated with increased intention to use STI-PrEP/PEP resulting from ordinal logistic regression among 524 MSM participating in the Amsterdam Cohort Study on HIV, the Netherlands, October 2021-April 2022.

## DISCUSSION

This study showed that a small fraction of MSM in the ACS currently use antibiotics to prevent STI. Awareness and use are highest among those using HIV-PrEP, had sexualized drug use or reported sexual behaviors with an increased chance of STI. Although preventive use of antibiotics was limited, given that a larger proportion had a high intention to use it, use may increase in the near future. This pattern was previously observed for HIV-PrEP in the same cohort (20,21).

Our data suggest that those who are more at risk for STI are more likely to be aware of biomedical STI prevention. Previously, a low intention for condom use related to willingness to engage in HIV-PrEP use (22,23) and a high incidence of STI among HIV-PrEP users has been reported (5–7). Just as with HIV-PrEP users, those intending to use STI-PrEP/PEP seek ways to reduce their chances of acquiring bacterial STI without hampering sexual pleasure by not using condoms. Condoms are often perceived to negatively affect sexual pleasure (24,25). For some, STI-PrEP/PEP might be a welcome approach to protect their health from frequent bacterial STI while having an active sexual life. For them, STI-PrEP/PEP may contribute to sexual wellbeing and quality of life. We also encountered first indications that STI-PrEP/PEP use is associated with stigma and that these perceptions are related to lower uptake intentions. Just like was the case with the early HIV-PrEP introduction.

An important additional finding in our study was that participants did not always use STI-PrEP/PEP adequately: they had used different types of antibiotics and different dosing schemes, which unintentionally can induce antimicrobial resistance and side effects. Hence, providing guidance and reliable information about STI-PrEP/PEP to MSM who informally use it, or are considering its use seems necessary.

Meanwhile, we need additional studies providing answers to the crucial questions that are still open. Closely monitoring resistance against doxycycline is important, as doxycycline is often a first-line and low-cost antibiotic in the treatment of sinusitis, infections with *Staphylococcus aureus* or *Coxiella burneti*, pneumonia, infections in people with amoxicillin hypersensitivity, amongst other indications. We need to be sure that doxy-PEP does not compromise its use as first-line medication to treat potentially life threatening infections and that its use does not jeopardize the World Health Organization (WHO) global action plan on AMR (26). Other potential drawbacks should be monitored as well, such as long-term effects on the microbiome, trends in condom norms and the spread of STIs that cannot be prevented with doxycycline.

The major strength of this study was its timeliness. The questions were introduced in an existing cohort at a time when this new strategy began to emerge as informal use despite the fact that the use of doxycycline or other antibiotics as prophylaxis against bacterial STI is not encouraged in the Netherlands. This study enabled us to measure the first practices, inclinations, and motives of the key population regarding this emerging strategy. A second strength was the heterogeneity of the study population regarding factors such as HIV status, HIV-PrEP use, drug use and sexual behavior. A limitation was that the numbers of actual users were too small for regression analyses. Since the ACS study population is a highly educated group actively involved in sexual health, and migrant populations are underrepresented, results may not be generalizable to the MSM population at large.

In conclusion, considering our results together with the results of clinical trials showing that the doxy-PEP effectively reduces STI risk (8–10), and the growing acceptance of biomedical prevention as witnessed for HIV-PrEP, we may expect that the demand for biomedical prevention for bacterial STI will increase in the future. In this light, we believe that close surveillance of actual STI-PrEP/PEP use is mandatory, together with research on the impact of doxy-PEP and STI-PrEP use on AMR. We need to develop a strategy to inform those who have already adopted STI-PrEP/PEP or have a high intention to do so, and their healthcare providers.

## Data Availability

All data produced in the present study are available upon reasonable request to the authors.

## ACKNOWLEDGEMENTS

The authors gratefully acknowledge the Amsterdam Cohort Studies (ACS) on HIV infection and AIDS, which is a collaboration between the Public Health Service of Amsterdam, Amsterdam UMC, Sanquin Blood Supply Foundation, Medical Center Jan van Goyen and the HIV Focus Center of the DC-Clinics. The authors wish to thank all ACS participants for their contribution, as well as the ACS study nurses, data managers and laboratory technicians.

## FUNDING

The Amsterdam Cohort Studies is part of the Netherlands HIV Monitoring Foundation and financially supported by the Center for Infectious Disease Control of the Netherlands National Institute for Public Health and the Environment, Bilthoven, the Netherlands. The authors declare no conflicts of interest.

## Notes

### Competing Interest Statement

The authors have declared no competing interest.

### Author Declarations

In 2022, the Amsterdam Cohort Studie on HIV was re-approved by the Medical Ethics Committee of the Amsterdam University Medical Center (UMC), the Netherlands (MEC 2007-182, NL18679.018.07).

## REFERENCES

1. Eisinger RW, Dieffenbach CW, Fauci AS. HIV viral load and transmissibility of HIV infection undetectable equals untransmittable. Vol. 321, JAMA - Journal of the American Medical Association. 2019.

2. Spinner CD, Boesecke C, Zink A, Jessen H, Stellbrink HJ, Rockstroh JK, et al. HIV pre-exposure prophylaxis (PrEP): a review of current knowledge of oral systemic HIV PrEP in humans. Vol. 44, Infection. 2016.

3. van Bilsen WPH, Boyd A, van der Loeff MFS, Davidovich U, Hogewoning A, van der Hoek L, et al. Diverging trends in incidence of HIV versus other sexually transmitted infections in HIV-negative MSM in Amsterdam. AIDS. 2020;34(2).

4. Unemo M, Bradshaw CS, Hocking JS, de Vries HJC, Francis SC, Mabey D, et al. Sexually transmitted infections: challenges ahead. Vol. 17, The Lancet Infectious Diseases. 2017.

5. Hoornenborg E, Coyer L, Boyd A, Achterbergh RCA, Schim van der Loeff MF, Bruisten S, et al. High incidence of HCV in HIV-negative men who have sex with men using pre-exposure prophylaxis. J Hepatol. 2020;72(5).

6. Hoornenborg E, Coyer L, van Laarhoven A, Achterbergh R, de Vries H, Prins M, et al. Change in sexual risk behaviour after 6 months of pre-exposure prophylaxis use: Results from the Amsterdam pre-exposure prophylaxis demonstration project. AIDS. 2018;32(11).

7. Traeger MW, Cornelisse VJ, Asselin J, Price B, Roth NJ, Willcox J, et al. Association of HIV Preexposure Prophylaxis With Incidence of Sexually Transmitted Infections Among Individuals at High Risk of HIV Infection. JAMA. 2019;321(14).

8. Molina JM, Charreau I, Chidiac C, Pialoux G, Cua E, Delaugerre C, et al. Post-exposure prophylaxis with doxycycline to prevent sexually transmitted infections in men who have sex with men: an open-label randomised substudy of the ANRS IPERGAY trial. Lancet Infect Dis. 2018;18(3):308–17.

9. Bolan RK, Beymer MR, Weiss RE, Flynn RP, Leibowitz AA, Klausner JD. Doxycycline prophylaxis to reduce incident syphilis among HIV-infected men who have sex with men who continue to engage in high risk sex: a randomized, controlled pilot study. Sex Transm Dis. 2015;42(2):98.

10. Luetkemeyer A, Donnell D, Dombrowski J, Cohen S, Grabow C, Brown C, et al. Postexposure doxycycline to prevent bacterial sexually transmitted infections. N Engl J Med. 2023;388(14):1296–306

11. San Francisco Department of Public Health. https://www.sfcdcp.org/wp-content/uploads/2022/10/Health-Update-Doxycycline-Post-Exposure-Prophylaxis-Reduces-Incidence-of-Sexually-Transmitted-Infections-SFDPH-FINAL-10.20.2022.pdf [Accessed on 24 February 2023]. 2022.

12. Lewis D. Push to use antibiotics to prevent sexually transmitted infections raises concerns. Nature. 2022;612:20–1.

13. Cars O, Mölstad S, Melander A. Variation in antibiotic use in the European Union. Lancet. 2001;357(9271).

14. Gradl G, Teichert M, Kieble M, Werning J, Schulz M. Comparing outpatient oral antibiotic use in Germany and the Netherlands from 2012 to 2016. Pharmacoepidemiol Drug Saf. 2018;27(12).

15. Two AM, Wu W, Gallo RL, Hata TR. Rosacea: Part II. Topical and systemic therapies in the treatment of rosacea. Vol. 72, Journal of the American Academy of Dermatology. 2015.

16. Zaenglein AL, Pathy AL, Schlosser BJ, Alikhan A, Baldwin HE, Berson DS, et al. Guidelines of care for the management of acne vulgaris. J Am Acad Dermatol. 2016;74(5).

17. Ajzen I. The theory of planned behavior. Organ Behav Hum Decis Process. 1991 Dec 1;50(2):179–211.

18. van Griensven GJ, de Vroome EM, Goudsmit J, Coutinho RA. Changes in sexual behaviour and the fall in incidence of HIV infection among homosexual men. Br Med J. 1989;298(6668):218–21.

19. SoaAids Nederland. https://www.soaaids.nl/files/2022-07/20220711-PrEP-richtlijn-Nederland-versie-3-update-2022.pdf [Accessed on 24 February 2023]. 2022.

20. Coyer L, van Bilsen W, Bil J, Davidovich U, Hoornenborg E, Prins M, et al. Pre-exposure prophylaxis among men who have sex with men in the Amsterdam Cohort Studies: Use, eligibility, and intention to use. PLoS One. 2018;13(10).

21. Bil JP, Davidovich U, van der Veldt WM, Prins M, de Vries HJC, Sonder GJB, et al. What do dutch msm think of preexposure prophylaxis to prevent HIV-infection? A cross study. AIDS. 2015;29(8).

22. Zimmermann HML, Jongen VW, Boyd A, Hoornenborg E, Prins M, de Vries HJC, et al. Decision-making regarding condom use among daily and event-driven users of preexposure prophylaxis in the Netherlands. AIDS. 2020;34(15).

23. Irungu EM, Ngure K, Mugwanya KK, Awuor M, Dollah A, Ongolly F, et al. “Now that PrEP is reducing the risk of transmission of HIV, why then do you still insist that we use condoms?” the condom quandary among PrEP users and health care providers in Kenya. AIDS Care - Psychological and Socio-Medical Aspects of AIDS/HIV. 2021;33(1).

24. Randolph ME, Pinkerton SD, Bogart LM, Cecil H, Abramson PR. Sexual pleasure and condom use. Arch Sex Behav. 2007;36(6).

25. Den Daas C, Adam PCG, Zuilhof W, de Wit JBF. A serological divide: men who have sex with men’s attitudes on HIV risk reduction strategies. AIDS Care - Psychological and Socio-Medical Aspects of AIDS/HIV. 2020;32(sup2).

26. World Health Organization (WHO). Global action plan on antimicrobial resistance. World Health Organization. 2017.

